# Pre-pandemic cross-reactive humoral immunity to SARS-CoV-2 in Africa: systematic review and meta-analysis

**DOI:** 10.1101/2022.10.07.22280814

**Authors:** John P.A. Ioannidis, Despina G. Contopoulos-Ioannidis

**Affiliations:** Departments of Medicine, of Epidemiology and Population Health, of Biomedical Data Sciences, and of Statistics, and Meta-Research Innovation Center at Stanford (METRICS), Stanford University, Stanford CA, USA; Division of Pediatric Infectious Diseases, Department of Pediatrics, Stanford University School of Medicine, Stanford, CA, USA

**Keywords:** COVID-19, SARS-CoV-2, malaria, cross-reactivity, pre-pandemic, seroprevalence

## Abstract

**Objective:** To assess the evidence on the presence of antibodies cross-reactive with SARS-CoV-2 antigens in pre-pandemic samples from African populations.

**Methods:** We performed a systematic review and meta-analysis of studies evaluating pre-pandemic African samples using pre-set assay-specific thresholds for SARS-CoV-2 seropositivity.

**Results:** 26 articles with 156 datasets were eligible, including 3,437 positives among 29,923 measurements (11.5%) with large between-dataset heterogeneity. Positivity was similar for anti-N (14%) and anti-S antibodies (11%), higher for anti-S1 (23%) and lower for anti-RBD antibodies (7%). Positivity was similar, on average, for IgM and IgG. Positivity was seen prominently in countries where malaria transmission occurs throughout and in datasets enriched in malaria cases (14%, 95% CI, 12-15% versus 2%, 95% CI 1-2% in other datasets). Substantial SARS-CoV-2 reactivity was seen in high malaria burden with or without high dengue burden (14% and 12%, respectively), and not without high malaria burden (2% and 0%, respectively). Lower SARS-CoV-2 cross-reactivity was seen in countries and cohorts of high HIV seroprevalence. More sparse individual-level data showed associations of higher SARS-CoV-2 cross-reactivity with *Plasmodium* parasitemia and lower SARS-CoV-2 cross-reactivity with HIV seropositivity.

**Conclusions:** Pre-pandemic samples from Africa show high levels of anti-SARS-CoV-2 seropositivity. Levels of cross-reactivity tracks especially with malaria prevalence.

## INTRODUCTION

Recorded COVID-19 deaths in Africa during the pandemic were few compared with other continents. Diverse explanations have been proposed [1], ranging from favorable population features (very young, low obesity rates) to undercounted deaths due to limited testing [2]. A most tantalizing hypothesis has invoked pre-existing immunity to SARS-CoV-2. Impetus for this hypothesis has been provided by studies assessing cross-reactive humoral immunity to SARS-CoV-2 in pre-pandemic African samples (Supplementary References [S1-S26]). This cross-reactivity has typically been absent in highly developed countries. Cross-reactivity to endemic coronaviruses is one possibility (e.g., see Supplementary References [S10, S25] for humoral and T-cell cross-reactivity), but it may not suffice to explain this phenomenon, and endemic coronaviruses are ubiquitously distributed across all continents. There have been speculations whether cross-reactive antibodies reflect exposure to other infectious pathogens widespread in Africa like *Plasmodium*, dengue and HIV.

African studies on cross-reactivity have used diverse sources of pre-pandemic samples in various countries. They have also used different assays for various antibody types and SARS-CoV-2 antigenic targets. There are conflicting hints on whether cross-reactivity may pertain mostly to specific antibody types and targets. Some studies (see Supplementary References [S2, S7, S8, S9, S10, S14, S15, S23, S25, S26) have also explored different correlations between the presence of anti-SARS-CoV-2 cross-reactivity and markers of various other pathogens. A systematic examination may offer some more concrete insights. Important questions to answer are: how frequently are SARS-CoV-2 cross-reactive antibodies detected in pre-pandemic African samples, how much heterogeneity exists on their prevalence, and whether heterogeneity may be explained by antibody types and targets, each country’s endemicity for *Plasmodium* and dengue, and HIV infection rates. This systematic review and meta-analysis addressed these questions.

## METHODS

The systematic review follows the PRISMA 2020 statement. The study protocol was registered on 9/13/2022 with registration number OSF f5g76 (https://osf.io/f5g76/).

### Eligible studies

Eligible studies were those that evaluated samples collected in African countries before December 2019, i.e. in the pre-pandemic period, for humoral responses to SARS-CoV-2 in blood (plasma or serum). Any type of antibody and any type of assay thereof was eligible. Studies were eligible if they used pre-set assay-specific thresholds of antibody titers for claiming positive results. Studies that measured antibodies in other body fluids (e.g., milk) were excluded. Studies that calibrated the assay threshold so as to make it appropriate for use in African populations by setting the specificity at a desired level were excluded; however, if these studies presented also specificity results according to an original pre-set threshold (based on previous work on other, non-African populations), the estimates of specificity based on the pre-set threshold were eligible. Similarly, studies that used assays that were previously calibrated so as to have a desirable level of specificity in African samples were excluded. Furthermore, studies that considered only pre-pandemic samples that were already screened to be negative for anti-SARS-CoV-2 antibodies based on some other SARS-CoV-2 antibody assay were excluded. For studies that included both African and other continent samples from the pre-pandemic era, only the former were eligible, and data were thus considered only if the African set could be separated. We did not consider functional (e.g. neutralizing antibody assays) or cellular immunity assays.

### Search strategies

PubMed was searched (last update May 11, 2023) with the following search string: (Nigeria OR Ethiopia OR Egypt OR Congo OR Tanzania OR South Africa OR Kenya OR Uganda OR Sudan OR Algeria OR Morocco OR Angola OR Mozambique OR Ghana OR Madagascar OR Cameroon OR Cote d’Ivoire OR Niger OR Burkina Faso OR Mali OR Malawi OR Zambia OR Senegal OR Chad OR Somalia OR Zimbabwe OR Guinea OR Rwanda OR Benin OR Burundi OR Tunisia OR South Sudan OR Togo OR Sierra Leone OR Libya OR Liberia OR Central African Republic OR Mauritania OR Eritrea OR Namibia OR Gambia OR Botswana OR Gabon OR Lesotho OR Guinea-Bissau OR Africa OR African) AND (pre-pandemic OR prepandemic OR cross-reactiv* OR seroprevalence OR negative samples OR negative controls OR (specificity AND test) OR (specificity AND antibod*)) AND (COVID-19 OR SARS-CoV-2). We also searched the reference lists of the retrieved eligible papers and searched in Google Scholar the articles that cite the retrieved eligible papers in order to identify any additional relevant eligible papers.

### Data extraction

From each eligible paper, we extracted the following information: first author, publication venue, African county(ies) from which pre-pandemic samples were obtained, sample size (per country and per cohort, if many countries/cohorts were assessed), provenance of the samples and any information about the sampling process, time periods when they were collected, age information; type of SARS-COV-2 assays and of antibodies measured, including manufacturer or laboratory of provenance, type of antibody (IgG, IgM, IgA, combinations, all antibodies) and antigenic targets (spike S, S1 subunit, S2 subunit, nucleocapsid N, receptor binding domain RBD, receptor binding motif RBM, other); number and percentage of positive samples for each antibody and antigenic target assessed among total assessed; any borderline readings; any additional information on measurements of indicators of other infectious diseases (*Plasmodium* parasitemia, *Plasmodium* antigens, HIV positivity status, dengue, other) with the potential to generate 2×2 tables for SARS-CoV-2 antibodies against these indicators; and any additional information on relationships between SARS-CoV-2 antibodies and other factors assessed by the authors.

Data extraction was performed in duplicate by two independent assessors who then compared notes and solved discrepancies with discussion.

### Risk of bias assessment

Eligible articles were assessed using the Joanna Briggs risk of bias tool for prevalence studies that includes 9 assessment items [3]. Assessment was done by one assessor.

### Meta-analysis

Available data on percentage of positive samples with different assays and antigenic targets were combined with meta-analysis using a random effects model [4]. Heterogeneity was assessed with the chi-square-based Q test and with the I^2^ statistic [5]. Borderline readings were considered negative (as also done by the original individual study authors).

Subgroup analyses were performed according to type of antibody (IgG, IgM, IgA, combinations, all antibodies). Moreover, when different types of antibody (e.g. IgG and IgM) had been assessed with the same assay platform in the same samples, the matched McNemar odds ratio was calculated.

We also performed subgroup analyses according to country of origin of the samples, classifying countries according to endemicity/burden for malaria, dengue, and rates of HIV infection. For malaria, we used the CDC map of malaria distribution [6] separating countries where malaria transmission occurs throughout versus those where no transmission occurs or transmission occurs only in some places. For dengue, we used the Global Consensus 2013 map [7] separating countries where dengue is present or likely from countries where dengue in uncertain, unlikely or absent. For HIV, we considered the eight countries with highest HIV positivity rates (>10% according to CIA fact book and UNAIDS AIDS info [8], i.e. Botswana, Lesotho, Mozambique, Namibia, South Africa, Eswatini, Zambia, and Zimbabwe) versus others. Studies where sampling explicitly aimed to recruit participants based on or heavily enriched in *Plasmodium*, dengue, or HIV infections were considered in the groups with high burden, even if they come from countries without high burden.

When many studies presented individual-level data to construct 2×2 tables for the association between anti-SARS-CoV-2 antibodies and other infectious disease markers, random effects meta-analyses of the calculated odds ratios were also performed. For the association between *Plasmodium* parasitemia and SARS-CoV-2 antibodies, we also used the Fisher method to combine p-values across studies that used different association metrics.

Meta-analyses were conducted in STATA SE 15. P-values were considered statistically significant for P<0.005 and suggestive for values between 0.005 and 0.05 [9].

## RESULTS

### Eligible studies

We screened 2,284 published items (Figure 1) of which 47 were screened in full text. 24 were excluded and 21 were eligible, while 3 more eligible studies (one article, one thesis and one preprint) were retrieved from cited/citation searches for a total of 26 eligible studies (Supplementary References [S1-26]). Studies screened in-depth and excluded are shown in Supplementary Excluded References along with reasons for exclusion. Supplementary Table 1 shows the results of the risk of bias assessment.

**Figure 1:**
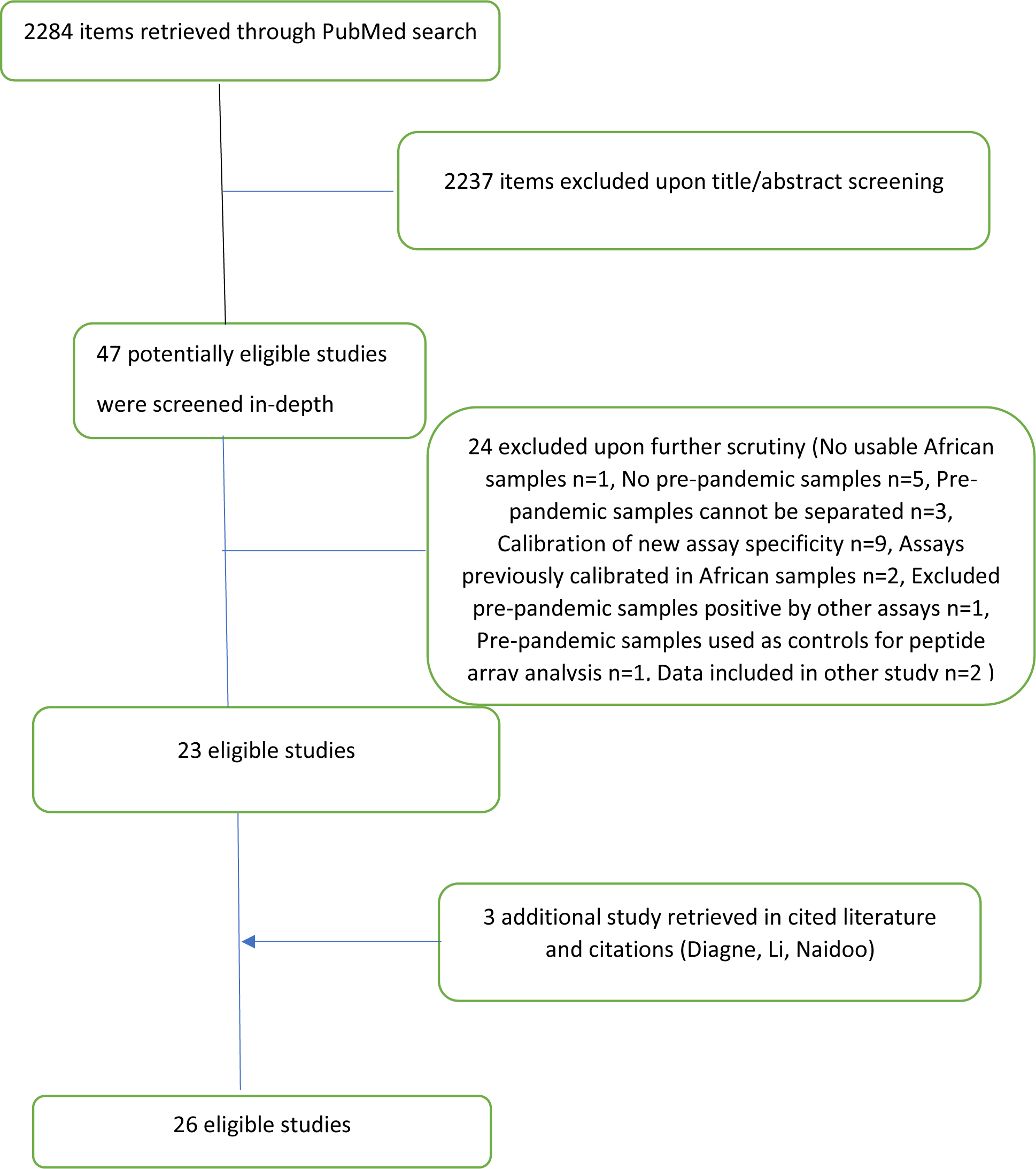
Flow diagram for searches and selected eligible studies

With one exception participants came from Sub-Saharan Africa (Table 1). Dataset sample sizes ranged from 19 to 1,077 (sum=29,923). There was also wide variation in the settings of sample collection and eligibility criteria. Six studies included independent cohorts from more than one country and two studies included different cohorts from the same country. Most samples were selected between 2016 and 2019, but 10 cohorts had earlier samples (earliest, 1999). Most cohorts had collected samples from both children and adults, but several included only adults and 2 included only children (Table 1).

**Table 1.** Characteristics of eligible studies

| **Author** | **Country** | **Sample size** | **Provenance of the samples and sampling process** | **Time period of sample collection** | **Age in years (median)** |
| --- | --- | --- | --- | --- | --- |
| Pedersen | Gabon | 146 | Unclear | Oct 2019 | 18-50 |
|  | Senegal | 150 | Unclear | Jan 2017-May 2018 | 65% <18 |
| Borrega | Sierra Leone | 120 | Lassa and Ebola survivors and their contacts | Sept 2016-April 2019 | 8-60 (31) |
| Tso | Tanzania | 105 | Blood donors, 6.7% HIV-positive | Mar 2019-May 2019 | ≥18 |
|  | Zambia | 99 | Enriched in HIV-positive (43.4%) | 2017- early<br>2019 | ≥18 |
| Emmerich | Magadascar | 167 | Pregnant women | 2010 | 20-30 (23) |
|  | Ghana | 150 | Children | 2014-2015 | 3-7 (6) |
|  | Ghana | 133 | Teens and adults | 1999 | 16-45 (22) |
|  | Nigeria | 150 | Adults | 2018 | 30-58 (41) |
| Yadouleton | Benin | 60 | Acute febrile illness, tested for hemorrhagic fever surveillance | Oct 2019 –<br>Nov 2019 | 12-65 (28) |
| Nzoghe | Gabon | 135 | Healthy healthcare worker volunteers | 2014 | 14-80 (38) |
| Woodford | Mali | 312 | Urban healthy adults | Jan 2017 | ≥18 |
|  |  |  | Rural healthy adults | May 2019 | ≥18 |
|  |  |  | Rural women of childbearing age | May 2019 | ≥18 |
|  |  |  | Rural all ages | May 2018 | All ages |
| Baker | Uganda | 1077 | Rakai Community Cohort Study (543 febrile within one month, 534 non-febrile matched for age and gender) | 2011-2013 | 18-54 (30) |
| Souris | DR Congo | 190 | Healthy subjects (hospital staff and volunteers) and young sickle-cell disease patients | 2019 | Any age |
|  | DR Congo | 384 | Biobank from Plasmodium study in Kinshasa | 2014 – 2015 | Any age |
|  | Cameroon | 383 | Continual health monitoring project for HIV patients | Jun 2018 – Jun 2019 | Any age |
|  | R Congo | 536 | Research samples from two districts (Brazaville, Bomassa) | 2016, 2019 | Any age |
|  | Senegal | 162 | Biobank samples | 2018, 2019 | Any age |
| Ige | Nigeria | 100 | 50 HBV-positive (S-antigen) and 50 HIV-positive | Before Oct 2019 | (35) |
| Traore | Mali | 283 | Malaria survey in Dangassa village | 2019 | Unclear |
| Ingoba | R Congo | 82 | Bomassa village | Jun- Jul 2019 | Unclear |
| Diagne | Senegal | 272 | Biobank | Before Sept 2019 | Unclear |
| Yansouni | Senegal | 100 | Clinical suspects for malaria | Before Jul 2019 | Unclear |
| Iriemenam | Nigeria 1 | 100 | Pre-pandemic samples of hepatitis B (n=50) or HIV (n=50) positive patients | Before Oct 2019 | (35) |
|  | Nigeria 2 | 200 | Nigeria HIV/AIDS Indicator and Impact Survey | 2018 | 0-60 (32) |
| Steinhardt | Nigeria | 213 | Nigeria HIV/AIDS Indicator and Impact Survey | 2018 | 0-60 |
| Gelanew | Ethiopia | 365 | Pre-pandemic sera | 2012-2018 | Unclear |
| Gebrecherkos | Ethiopia | 50 | Pre-pandemic samples from patients with other infections | 2017 | Unclear |
| Mboumba<br>Bouassa | Central<br>African<br>Republic | 100 | National center of sexually transmitted diseases in Bangui, 54 HIV- seropositive, 5 HCV-positive, 35 HBsAg-positive | 2000-2011 | (31 mean) |
| Gdoura | Tunisia | 119 | Pre-pandemic samples | Before 2019 | Unclear |
| Vanroye | Diverse African | 195<sup>a</sup> | Travelers from Africa with malaria or schistosomiasis | 2010-2018 | 8-61 (~40) |
| Lapidus | Cameroon | 19 | Malaria patients | Jul-Nov 2018 | 2-64 (26 mean) |
|  | Senegal 1 | 120 | Malaria patients | Jul 2019 | 1-74 (22 mean) |
|  | Senegal 2 | 67 | Malaria patients | 2015-2017 | 5-16 (11 mean) |
|  | Burkina Faso 1 | 88 | Malaria patients | Jul-Aug 2017 | 0-4 (3 mean) |
|  | Burkina Faso 2 | 25 | Malaria patients | Oct 2016-Feb 2017 | 21-43 (33) |
|  | Ghana | 45 | Malaria patients | Jul 2007-June 2010 | 3-70 (15) |
| Naidoo | South Africa | 316 | Pre-pandemic virology serology residual samples | Before November 2019 | Unclear |
| Li | Nigeria | 361 | RV466/IWARG (febrile illness study) | 2014-2019 | 18-78 (34) |
|  | Uganda | 399 | RV329/AFRICOS (mostly chronic HIV infection) | 2014-2016 | 20-70 (40) |
|  | Tanzania | 234 | RV329/AFRICOS (mostly chronic HIV infection) | 2014-2016 | 19-74 (39) |
|  | Kenya | 653 | RV329/AFRICOS (mostly chronic HIV infection) | 2014-2017 | 19-77 (40) |
| Nantambi | Uganda | 110 | Specimens collected for optimization of immunological assays, 68% HIV-positive | 2009-1015 | Unclear |
| Aissatou | Cameroon | 288 | Biobank samples from Centre region of Cameroon, 56% HIV-positive | 2017-1018 | 0=>60 (25) |

<sup>a</sup>Includes 9 samples from Asia and 19 of unknown country origin; besides the n=195, the study has 25 patients with dengue, but few are from Africa, so only the malaria and schistosomiasis cases are considered here

There was a large variety of assays used, but most studies used assays of IgG (Table 2). Most studies assessed antibodies against both spike and nucleocapsid, but antigenic targets varied across studies (Table 2). In-depth assessments for associations with indicators of infectious pathogens were sparse.

**Table 2.** Immunological assays performed in the eligible studies and other pathogen indicators assessed

| Author | Antibody assays used | Antigenic targets | Other pathogen indicators assessed |
| --- | --- | --- | --- |
| Pedersen | ELISA | S, N |  |
| Borrega | reSARS, cutoffs defined by USA samples | N, RBD, S2 |  |
| Tso | Immunofluorescence assay, with USA control samples | N, S | HIV |
| Emmerich | Euroimmun, EDI, Mikrogen recomWell | S1 IgG (Euroimmun), N IgG (Euroimmun), N IgG (EDI), N IgG (Mikrogen) | *Plasmodium* parasitemia |
| Yadouleton | Euroimmun, INBIOS | S IgG (Euroimmun), S1 IgA (Euroimmun), S1 IgG (INBIOS), N IgG (Euroimmun) |  |
| Nzoghe | Elecsys Roche | N all subclasses | Multiple infectious indicators assessed but presented only for SARS-C0V-2 antibody-positive samples |
| Woodford | Previously developed at NIH with USA samples | S, N, RBD | *Plasmodium* antigens |
| Baker | CoronaCHEK | Spike RBD IgG or IgM | HIV |
| Souris | INNOBIOCHIPS ELISA, calibrated on other human coronaviruses | S1, S2, S1-RBD, S1-NTD, N |  |
| Ige | Euroimmun, Mologic, Abbott Architect | S1 IgG (Euroimmun), N IgG (Euroimmun), S2 or N IgG (Mologic), N IgG (Abbott) |  |
| Traore | ELISA | S IgG, RBD IgG, RBM IgG | *Plasmodium* parasitemia, *Plasmodium* smear |
| Ingoba | Virotech, according to kit manufacturer | N IgG (also IgM but no data given) |  |
| Diagne | Omega, ID-screen | N or S2 IgG (Omega), N IgG (ID-screen) | *Plasmodium* and other pathogen antibodies |
| Yansouni | Abbott Architect, Rapid diagnostic test (RDT) | N IgG (Abbott), N IgM (RDT), N IgG (RDT) |  |
| Iriemenam | Tetracore, SARS2MBA, xMAP, RightSign | RBD IgG, N IgG, RBD-N hubrid IgG (Tetracore); S IgG, RBD541 IgG, |  |
|  |  | RBD591 IgG, N IgG<br>(SARS2MBA); N IgG,<br>RBD IgG, S1 IgG<br>(xMAP); IgG/IgM<br>(RightSign) |  |
| Steinhardt | Euroimmun, Abbott<br>Architect | N IgG (Euroimmun), N<br>IgG (Abbott) | *Plasmodium* antibodies,<br>*Plasmodium* antigens,<br>antibodies for filariasis,<br>oncocherciasis,<br>syphilis/yaws,<br>cystocercosis, taeniasis |
| Gelanew | ELISA | RBD IgG |  |
| Gebrecherkos | Canea, Cellex, VivaChek,<br>Innovita; ECLIA Roche | N+S IgG or IgM (Cenea,<br>Cellex, VivaChek,<br>Innovite), N all<br>immunoglobulin (Roche) |  |
| Mboumba<br>Bouassa | BIOSYNEX, SIENNA, NG-test | RBD IgG and RBD IgM<br>(BIOSYNEX), unknown<br>antigen IgG and unknown<br>antigen IgM (SIENNA),<br>unknown antigen IgG and unknown antigen IgM<br>(NG-test) |  |
| Gdoura | Vidas Biomerieux, Mindray,<br>Elecsys Roche (Cobas),<br>Access | S1 RBD IgG and S1 IgM<br>(Vidas), S+N IgG and<br>S+N IgG (Mindray), N all<br>subclasses (Elecsys), S1<br>RBD IgG (Access) |  |
| Vanroye | 13 RDTs | N IgG or IgM (Toda,<br>Biohit, Panbio, Boson),<br>N+S IgG or IgM (Cellex,<br>Dynamiker, Liming Bio),<br>RBD IgG or IgM<br>(ZenTech), N+RBD IgG<br>or IgM (SureScreen<br>Diagnostics, Singuway),<br>N+RBD+S IgG or IgM<br>(Multi-G), S IgG or IgM<br>(Healgen), S (Wantai) |  |
| Lapidus | ELISA | S1 IgG, S1 IgM | All participants have<br>had malaria (different<br>clinical phenotypes) |
| Naidoo | Elecsys Roche | N all subclasses, S all subclasses |  |
| Li | Bead-based assay | S2 IgG | HIV, *Plasmodium* smear (no statistically significant differences); several others, but only in those samples with anti-S2 reactivity |
| Nantambi | In-house ELISA | N IgG, S IgG | HIV |
| Aissatou | Panbio IgG, IgM, IgG/IgM | N IgG, N IgM, N IgG/IgM | HIV |

### Presence of SARS-CoV-2 antibodies

Overall, 156 datasets (Supplementary Table 2) among pre-pandemic samples were available for analysis with crude total of 3,437 positives among 29,923 measurements (11.7%, 95% CI, 11.1-11.9%). When all 156 datasets were considered in meta-analysis, there was large between-dataset heterogeneity (p<0.001, I^2^=96.5%) with several studies finding 0% positivity and some others exceeding 80% (summary random effects 11%, 95% CI, 10-12%).

### Meta-analysis of positivity per antigenic target and antibody type

As shown in Table 3, summary positivity was similar for anti-N (14%, 95% CI, 11-16%) and anti-S (11%, 95% CI, 10-12%) antibodies. However, subtypes within spike had different profiles and summary positivity was higher for anti-S1 (23%, 95% CI, 18-27%) and lower for anti-RBD antibodies (7%, 95% CI, 5-8%). Lowest positivity was seen for antibodies using both N and S (or subtype) targets (5%, 95% CI, 3-7%). Positivity was overall similar for IgM (summary 13%) and IgG (summary 11%) antibodies, but prominence of IgM versus IgG signals varied greatly across datasets. There was very large between-dataset heterogeneity in all summary estimates (p<0.001), so they should be seen with great caution.

**Table 3.**
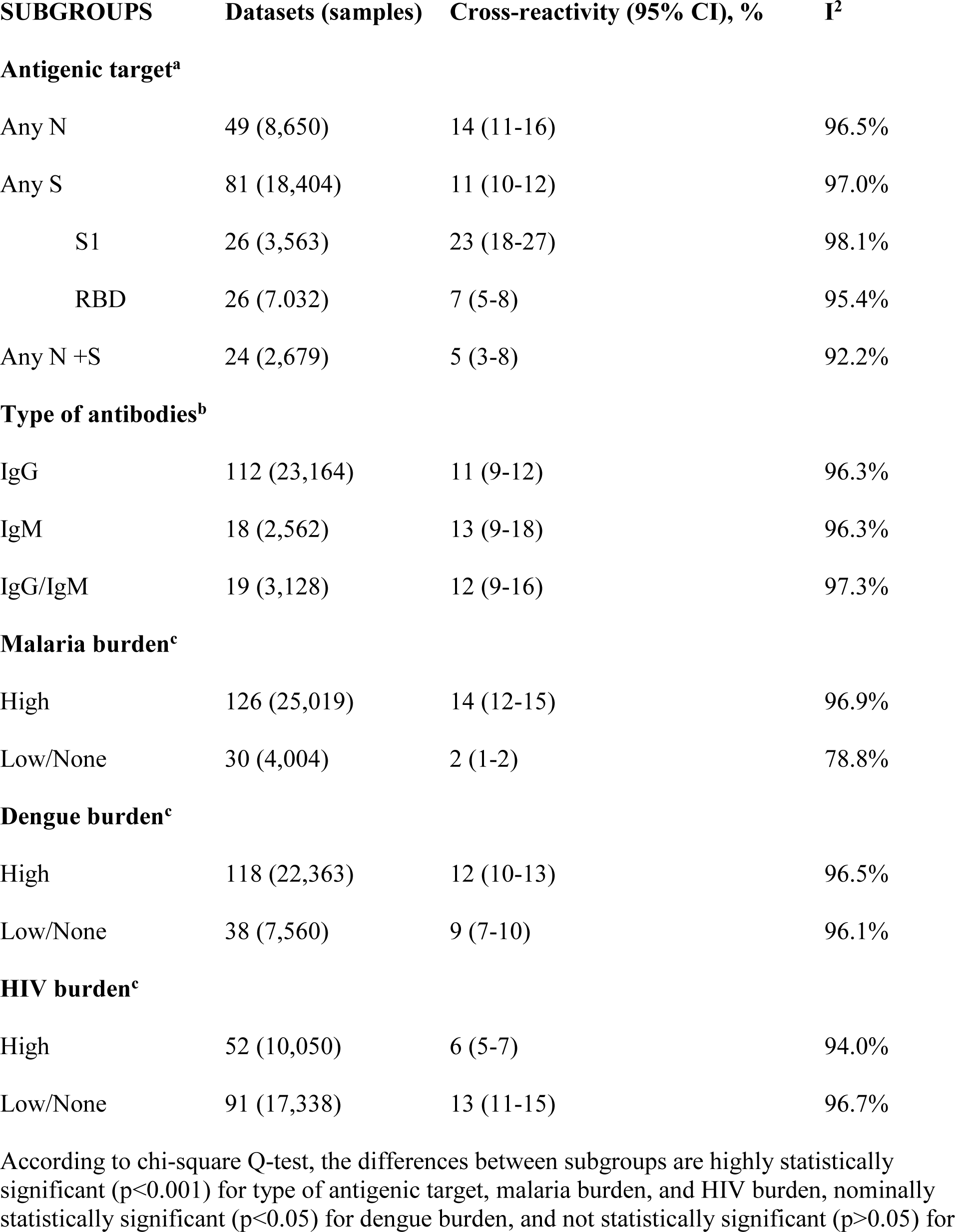
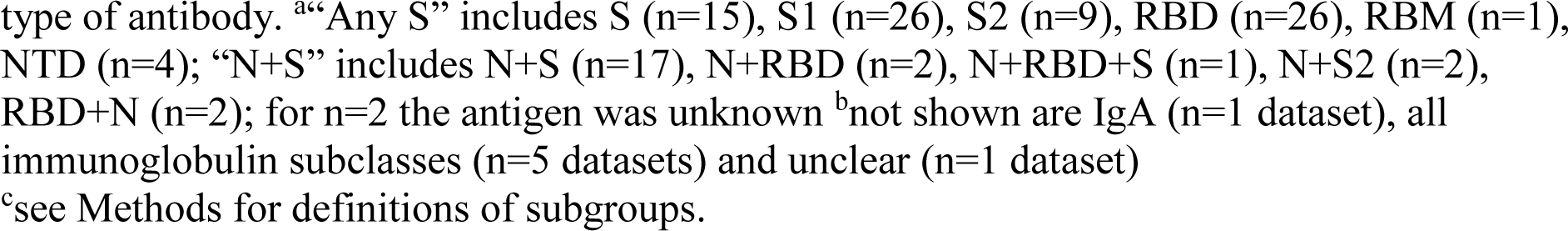
Summary estimates of anti-SARS-CoV-2 cross-reactivity

| **SUBGROUPS** | **Datasets (samples)** | **Cross-reactivity (95% CI), %** | **I<sup>2</sup>** |
| --- | --- | --- | --- |
| **Antigenic target<sup>a</sup>** |  |  |  |
| Any N | 49 (8,650) | 14 (11-16) | 96.5% |
| Any S | 81 (18,404) | 11 (10-12) | 97.0% |
| S1 | 26 (3,563) | 23 (18-27) | 98.1% |
| RBD | 26 (7,032) | 7 (5-8) | 95.4% |
| Any N +S | 24 (2,679) | 5 (3-8) | 92.2% |
| **Type of antibodies<sup>b</sup>** |  |  |  |
| IgG | 112 (23,164) | 11 (9-12) | 96.3% |
| IgM | 18 (2,562) | 13 (9-18) | 96.3% |
| IgG/IgM | 19 (3,128) | 12 (9-16) | 97.3% |
| **Malaria burden<sup>c</sup>** |  |  |  |
| High | 126 (25,019) | 14 (12-15) | 96.9% |
| Low/None | 30 (4,004) | 2 (1-2) | 78.8% |
| **Dengue burden<sup>c</sup>** |  |  |  |
| High | 118 (22,363) | 12 (10-13) | 96.5% |
| Low/None | 38 (7,560) | 9 (7-10) | 96.1% |
| **HIV burden<sup>c</sup>** |  |  |  |
| High | 52 (10,050) | 6 (5-7) | 94.0% |
| Low/None | 91 (17,338) | 13 (11-15) | 96.7% |
| According to chi-square Q-test, the differences between subgroups are highly statistically significant (p<0.001) for type of antigenic target, malaria burden, and HIV burden, nominally statistically significant (p<0.05) for dengue burden, and not statistically significant (p>0.05) for |  |  |  |

type of antibody. <sup>a</sup>“Any S” includes S (n=15), S1 (n=26), S2 (n=9), RBD (n=26), RBM (n=1), NTD (n=4); “N+S” includes N+S (n=17), N+RBD (n=2), N+RBD+S (n=1), N+S2 (n=2), RBD+N (n=2); for n=2 the antigen was unknown <sup>b</sup>not shown are IgA (n=1 dataset), all immunoglobulin subclasses (n=5 datasets) and unclear (n=1 dataset)
<sup>c</sup>see Methods for definitions of subgroups.

### Paired comparisons of antibody types

In 18 datasets, both IgG and IgM had been measured with the same type of assays and antigenic target on the same samples. For 16/18, data on paired measurements in each sample were available (Supplementary Table 3, in another 2 small datasets had no sufficient data to calculate paired odds ratios). The results were very heterogeneous making their synthesis precarious: in 7 datasets 95% confidence intervals of the paired odds ratio excluded the null, but in 4 datasets, samples positive only by IgM outnumbered samples positive only by IgG, while the opposite situation was seen for 3 datasets.

### Subgroup analyses for malaria, dengue, and HIV (Table 3)

Anti-SARS-CoV-2 positivity was very different according to malaria status. It was seen prominently in countries where malaria transmission occurs throughout and in datasets enriched in malaria cases (summary estimate 14%, 95% CI, 12-15%), but was almost absent in other datasets (summary estimate 2%, 95% CI, 1-2%). There was still very large between-study heterogeneity in the former group, and less heterogeneity in the latter. Among the 30 datasets in the latter group, positivity was always 0% or very low (=<8%) except for one dataset from Tanzania which is a country where malaria transmission does occur throughout in altitudes below 1800 meters.

A higher proportion of anti-SARS-CoV-2 positivity was seen in high dengue burden datasets than others (summary estimates 12% versus 9%), but large between-dataset heterogeneity existed within each group (p<0.001).

When data were analyzed in 4 subgroups according to both malaria and dengue burden (Supplementary Figure), seropositivity was similar in settings with high malaria burden regardless of whether they had high or low dengue burden (14% [95% CI, 13-16%] and 12% [95% CI, 9-14%], respectively). Conversely, there was low or no seropositivity in settings with low malaria burden, regardless of whether they had high or low dengue burden (2% [95% CI, 1-3%] and 0% [95% CI, 0-1%], respectively).

An inverse association was seen with HIV infection, with lower proportions of anti-SARS-CoV-2 antibody positivity in countries with high HIV transmission or datasets with >10% of the sample being HIV-positive than in datasets with lower HIV rates (summary positivity 6% versus 13%). There was large between-dataset heterogeneity (p<0.001) within each group.

### Association with Plasmodium parasitemia (data at sample level)

One study (supplementary reference [S9]) provided 2×2 tabular data on positivity with 4 different anti-SARS-CoV-2 IgG assays in subgroups defined by the presence of *Plasmodium* parasitemia. Combining the 4 evaluations, summary odds ratio was 1.84 (95% CI, 0.90-3.78, I^2^=29.2%). 15 evaluations from 3 studies (supplementary references [S2, S8, S9]) presented various examinations of the association between *Plasmodium* parasitemia and anti-SARS-CoV-2 cross-reactivity using diverse metrics: combining the 15 p-values yielded p=0.009 for the overall association (Supplementary Table 4).

### Association with HIV seropositivity (data at sample level)

Four studies (supplementary references [S7, S10, S25, S26]) included data from 8 datasets where SARS-CoV-2 cross-reactivity was given per HIV serostatus (Supplementary Table 5). Summary odds ratio for the association between SARS-CoV-2 antibodies and HIV seropositivity was 0.69 (0.49-0.96, I^2^=0%).

Data were limited or not presented in sufficient detail for formal meta-analysis for other infectious disease indicators.

## DISCUSSION

The present meta-analysis includes data from 26 studies with 156 datasets and approximately 30,000 measurements of anti-SARS-CoV-2 antibodies in pre-pandemic samples from Africa. On average, approximately 1 of 9 samples tested positive for anti-SARS-CoV-2 antibodies, but there was extensive heterogeneity across studies and datasets. Stark differences were seen according to malaria burden. Anti-SARS-CoV-2 cross-reactivity was seen almost entirely in samples from areas with malaria transmission throughout and/or enriched in malaria cases. A more modest association was seen for dengue. However, malaria and dengue endemicity largely overlap, and samples coming from high dengue but low malaria burden settings had negligible anti-SARS-CoV-2 cross-reactivity. Finally, HIV was associated with lower frequency of anti-SARS-CoV-2 antibodies. These associations with pathogen burden are based on inferences about the frequency of these infections at group/country-level rather than individual samples; therefore they should be seen with great caution for potential ecological fallacies. Nevertheless, also in the more sparse data on individual samples, *Plasmodium* parasitemia was associated with higher SARS-CoV-2 cross-reactivity and HIV seropositivity was associated with lower SARS-CoV-2 cross-reactivity.

The composite picture is consistent with the possibility that the observed pre-pandemic seropositivity to SARS-CoV-2 in Africa may partly reflect cross-reactive response to malaria. Lapidus et al. suggest that this immune response is more common and more intense in acute and recent malaria [10], may reflect antibodies targeting sialic acid moieties on the S1 fragment, and cross-reactivity is effectively reduced when sialic acid is cleaved from the assay target [10]. The large between-dataset heterogeneity among studies with high malaria burden may reflect both antibody assay variability and diversity in the magnitude of the malaria burden: some datasets included exclusively acute malaria, others were heavily enriched in malaria cases, and others simply came from areas with substantial malaria burden. Most areas in Africa with high dengue burden have also high malaria burden. Datasets from settings with high dengue but low malaria showed negligible anti-SARS-CoV-2 positivity. Prospective studies with pandemic and post-pandemic samples should further evaluate the relationship between malaria and SARS-CoV-2 seropositivity.

Anti-SARS-CoV-2 humoral cross-reactivity in pre-pandemic samples has generally not been observed in European and USA studies. Data from countries outside Africa also suggest that malaria rather than dengue has a strong association with SARS-CoV-2 cross-reactive antibodies. Manning et al. [11] found 14% cross-reactive positivity among 528 malaria patients from Cambodia. More studies on individual samples with documented malaria are needed from non-African areas. Data for dengue are mixed. One study in Taiwan [12] found higher optical density anti-S1 RBD activity in archival dengue samples than controls, but optical density values were still low. Another study [13] found some IgM and IgA rather IgG false-positivity for SARS-CoV-2 in dengue febrile illness in Thailand, but false-positivity tended to be even more frequent for non-dengue febrile illness (including apparently malaria). In samples from Puerto Rico and USA [14], dengue did not induce cross-reactive antibodies to SARS-CoV-2; the same was true in dengue samples from Indonesia [15], Colombia [16] and travel clinics [17]. Conversely, another study [18] found 22% cross-reactivity in samples from an Israel center (unspecified country of provenance); however, it is unknown whether any positive samples could be from patients with malaria history. SARS-CoV-2 infection produces cross-reactive antibody activity to dengue in some [14,15,18] but not all [13] studies. Anti-SARS-CoV-2 antibodies may also have a protective role for dengue [11]. Interestingly, reported dengue cases and deaths declined in 2020-2022 after a peak in 2019 [19], but this may also reflect the impact of containment measures on communicable diseases in general. In-silico analysis shows possible similarities between SARS-CoV-2 epitopes in the HR2 domain of spike and the dengue envelope protein [18], but evidence is again stronger for malaria, where cross-reactive antibodies specifically recognized the sialic acid moiety on N-linked glycans of spike [10].

The clinical and public health importance of pre-existing humoral cross-reactivity remains a tantalizing question. Evidence is mixed. Some studies have suggested that the detected antibodies in pre-pandemic samples test negative in neutralization assays [10,20]. Conversely, Li et al. [21] found neutralizing responses in some pre-pandemic samples; neutralizing and binding activities did not correlate. Stronger evidence for neutralizing activity was obtained by Nyagwange et al [22]. who analyzed pre-pandemic samples from coastal Kenya and found neutralization for pseudotyped reference SARS-CoV-2 with a mean infective dose 50 of 1:251, which is still within predicted protection levels. In the same study, the pre-pandemic naso-oropharyngeal fluid samples also neutralized pseudo-SARS-CoV-2 at a mean infective dose 50 of 1:5.9.

These cross-reactive antibodies may be a marker of a much broader immune response with both humoral and cellular features. Pre-existing T-cell immunity and its potential role in ameliorating clinical course in SARS-CoV-2 infection is another debated issue [23]. It would be useful to assess pre-pandemic samples with anti-SARS-CoV-2 antibodies for a broad spectrum of immune functionalities. Non-humoral immunity elements appear even more frequent than the detected humoral immunity, since humoral immunity tends to wane relatively rapidly [24]. Cross-reactive CD4+ and CD8+ T-cell responses to SARS-CoV-2 were documented in 90% of pre-pandemic samples from Uganda [25], but only 10-50% in pre-pandemic samples from the USA or Europe [26–28]. Cross-reactive T-cell responses may derive from both homologous pathogens (other coronaviruses, including the 4 globally-distributed coronaviruses or other coronaviruses with country distribution largely overlapping with malaria burden [20]) and non-homologous ones (e.g. CMV) [29].

The geographical pattern of the documented impact of COVID-19 in Africa is intriguingly well-aligned with the geographical pattern of detected pre-pandemic humoral cross-reactivity. Recorded COVID-19 deaths have been far higher in South Africa (high HIV, relatively low malaria burden) and in northern African countries (low malaria burden) than in other countries in sub-Saharan Africa (high malaria burden). These areas are unlikely to have differed in prior exposure to the 4 well-known endemic coronaviruses. However, differences in other environmental conditions or in the extent of contact with wildlife carrying other coronaviruses are an alternative possibility [20]. Differences in the extent of under-ascertainment of COVID-19 deaths, demographic and lifestyle differences (older populations in northern Africa, high levels of obesity in South Africa) may also partly underlie these differences. However, a contribution of pre-existing immunity remains also an additional possibility. Pre-existing immunity may have contributed also to relatively low fatalities in East Asia. Many areas in East Asia also have substantial malaria burden. Conversely, recent dengue outbreaks did not seem to protect from COVID-19 fatalities; the highest dengue burden in 2019 was seen in Brazil, a country that suffered high COVID-19 fatalities.

Our work has limitations. First, the examined studies mostly used convenient samples available from pre-pandemic efforts not tailored specifically to answer questions posed by the pandemic. For many samples, information about their provenance and features was limited. Second, several analyses have ecological designs, e.g. when countries were assigned to high or low burden groups for specific pathogens. The observed associations may not necessarily hold true also in analyses profiling prior infection in single individual samples. The potential for ecological fallacy plus the high between-dataset heterogeneity should add extra caution to the interpretation of the results of the presented meta-analyses. Nevertheless, the more limited individual sample-level data available also agree with the main findings regarding malaria [30] and HIV. Third, some analyses include datasets which represent the same samples tested with different assays, therefore they are not entirely independent. However, the major differences observed (e.g. with malaria) remain strong even if only one dataset is selected per study/cohort (not shown). Fourth, it is uncertain if due to publication bias more studies that found no seropositivity in pre-pandemic African samples may have remained unpublished compared with studies that found high positivity. Fifth, the assays used were very diverse and the technical competence of their performance by different teams cannot be validated independently. This may explain also part of the observed large between-dataset heterogeneity. However, errors would tend to weaken observed associations, if anything, through non-differential mis-classification.

Acknowledging these caveats, our meta-analysis provides strong evidence for pre-pandemic cross-reactive humoral immunity to SARS-CoV-2 in Africa, closing tracking with malaria. Further studies of broader immunological profiles involved and of the public health implications are necessary.

## Data Availability

All data are in the manuscript and in its supplements

## Contributors

Both authors conceived the original idea, wrote the protocol, extracted data, run and interpreted analyses, wrote and revised the paper and approved the final version.

## Data sharing statement

All data are in the manuscript and its supplements

## Ethical approval

Not applicable

