## Supplementary Appendix file for "Pre-pandemic cross-reactive humoral immunity to SARS-CoV-2 in Africa: systematic review and meta-analysis"

**Supplementary References S1-26**

1. Pedersen J, Koumakpayi IH, Babuadze G, et al. Cross-reactive immunity against SARS-CoV-2 N protein in Central and West Africa precedes the COVID-19 pandemic. Sci Rep. 2022 Jul 28;12(1):12962.
2. Traoré A, Guindo MA, Konaté D, et al. Seroreactivity of the Severe Acute Respiratory Syndrome Coronavirus 2 Recombinant S Protein, Receptor-Binding Domain, and Its Receptor-Binding Motif in COVID-19 Patients and Their Cross-Reactivity With Pre-COVID-19 Samples From Malaria-Endemic Areas. Front Immunol. 2022 Apr 27;13:856033.
3. Iriemenam NC, Ige FA, Greby SM, Okunoye OO, Uwandu M, Aniedobe M, Nwaiwu SO, Mba N, Okoli M, William NE, Ehoche A, Mpamugo A, Mitchell A, Stafford KA, Thomas AN, Olaleye T, Akinmulero OO, Agala NP, Abubakar AG, Owens A, Gwyn SE, Rogier E, Udhayakumar V, Steinhardt LC, Martin DL, Okoye MI, Audu R. Comparison of one single-antigen assay and three multi-antigen SARS-CoV-2 IgG assays in Nigeria. J Clin Virol Plus. 2023 Feb;3(1):100139.
4. Borrega R, Nelson DKS, Koval AP, et al. Cross-Reactive Antibodies to SARS-CoV-2 and MERS-CoV in Pre-COVID-19 Blood Samples from Sierra Leoneans. Viruses. 2021 Nov 21;13(11):2325.
5. Ige F, Hamada Y, Steinhardt L, et al. Validation of Commercial SARS-CoV-2 Immunoassays in a Nigerian Population. Microbiol Spectr. 2021 Oct 31;9(2):e0068021.
6. Woodford J, Sagara I, Dicko A, et al. Severe Acute Respiratory Syndrome Coronavirus 2 Seroassay Performance and Optimization in a Population With High Background Reactivity in Mali. J Infect Dis. 2021 Dec 15;224(12):2001-2009.
7. Baker OR, Grabowski MK, Galiwango RM, et al. Differential Performance of CoronaCHEK SARS-CoV-2 Lateral Flow Antibody Assay by Geographic Origin of Samples. J Clin Microbiol. 2021 Jun 18;59(7):e0083721.
8. Steinhardt LC, Ige F, Iriemenam NC, et al. Cross-Reactivity of Two SARS-CoV-2 Serological Assays in a Setting Where Malaria Is Endemic. J Clin Microbiol. 2021 Jun 18;59(7):e0051421.
9. Emmerich P, Murawski C, Ehmen C, et al. Limited specificity of commercially available SARS-CoV-2 IgG ELISAs in serum samples of African origin. Trop Med Int Health. 2021 Jun;26(6):621-631.
10. Tso FY, Lidenge SJ, Peña PB, et al. High prevalence of pre-existing serological cross-reactivity against severe acute respiratory syndrome coronavirus-2 (SARS-CoV-2) in sub-Saharan Africa. Int J Infect Dis. 2021 Jan;102:577-583.
11. Yansouni CP, Papenburg J, Cheng MP, et al. Specificity of SARS-CoV-2 Antibody Detection Assays against S and N Proteins among Pre-COVID-19 Sera from Patients with Protozoan and Helminth Parasitic Infections. J Clin Microbiol. 2022 Jan 19;60(1):e0171721.
12. Lapidus S, Liu F, Casanovas-Massana A, Dai Y, Huck JD, Lucas C, Klein J, et al. Plasmodium infection is associated with cross-reactive antibodies to carbohydrate epitopes on the SARS-CoV-2 spike protein. Sci Rep. 2022;12(1):22175.
13. Yadouleton A, Sander AL, Moreira-Soto A, et al. Limited Specificity of Serologic Tests for SARS-CoV-2 Antibody Detection, Benin. Emerg Infect Dis. 2021 Jan;27(1):233–7.
14. Diagne CT, Ndiaye O, Talla C, et al. Cross-Reactivity of SARS-CoV-2 Laboratory Diagnostics to Endemic Diseases in Africa: A Diagnostic Accuracy Study. Available at SSRN: http://dx.doi.org/10.2139/ssrn.3916756
15. Mveang Nzoghe A, Essone PN, Leboueny M, et al. Evidence and implications of pre-existing humoral cross-reactive immunity to SARS-CoV-2. Immun Inflamm Dis. 2021 Mar;9(1):128-133.
16. Souris M, Tshilolo L, Parzy D, Lobaloba Ingoba L, Ntoumi F, Kamgaing R, Ndour M, Mbongi D, Phoba B, Tshilolo MA, Mbungu R, Sosso MS, Fainguem N, Ndiaye Dieye T, Sylla M, Morand P, Gonzalez JP. Pre-Pandemic cross-reactive immunity against SARS-CoV-2 among Central and West African populations. Viruses. 2022 Oct 14;14(10):2259.
17. Lobaloba Ingoba L, Djontu JC, Mfoutou Mapanguy CC, et al. Seroprevalence of anti-SARS-CoV-2 antibodies in a population living in Bomassa village, Republic of Congo. IJID Reg. 2022 Mar;2:130-136.
18. Gdoura M, Halouani H, Sahli D, Mrad M, Chamsa W, Mabrouk M, Hogga N, Ben-Salem K, Triki H. SARS-CoV-2 serology: utility and limits of different antigen-based tests through the evaluation and the comparison of four commercial tests. Biomedicines. 2022 Dec 1;10(12):3106.
19. Gebrecherkos T, Kiros YK, Challa F, et al. Longitudinal profile of antibody response to SARS-CoV-2 in patients with COVID-19 in a setting from Sub-Saharan Africa: A prospective longitudinal study. PLoS One. 2022 Mar 23;17(3):e0263627.
20. Gelanew T, Seyoum B, Mulu A, et al. High seroprevalence of anti-SARS-CoV-2 antibodies among Ethiopian healthcare workers. BMC Infect Dis. 2022 Mar 16;22(1):261.
21. Mboumba Bouassa RS, Péré H, Tonen-Wolyec S, et al. Unexpected high frequency of unspecific reactivities by testing pre-epidemic blood specimens from Europe and Africa with SARS-CoV-2 IgG-IgM antibody rapid tests points to IgM as the Achilles heel. J Med Virol. 2021 Apr;93(4):2196-2203.
22. Vanroye F, Bossche DVD, Brosius I, Tack B, Esbroeck MV, Jacobs J. COVID-19 Antibody Detecting Rapid Diagnostic Tests Show High Cross-Reactivity When Challenged with Pre-Pandemic Malaria, Schistosomiasis and Dengue Samples. Diagnostics (Basel). 2021 Jun 25;11(7):1163.
23. Li Y, Merbah M, Wollen-Roberts S, Beckman B, Mdluli T, Swafford I, Mayer SV, King J, Corbitt C, Currier JR, Liu H, Esber A, Pinyakorn S, Parikh A, Francisco LV, Phanuphak N, Maswai J, Owuoth J, Kibuuka H, Iroezindu M, Bahemana E, Vasan S, Ake JA, Modjarrad K, Gromowski G, Paquin-Proulx D, Rolland M. Coronavirus Antibody Responses before COVID-19 Pandemic, Africa and Thailand. Emerg Infect Dis. 2022 Nov;28(11):2214-2225.
24. Naidoo M. Evaluation of Two SARS-CoV-2 Immunoassays, thesis NDXMIC018, submitted to the University of Cape Town, 2022, in: <https://open.uct.ac.za/handle/11427/37603>, last accessed April 4, 2023.
25. Nantambi H, Sembera J, Ankunda V, Ssali I, Kalyebi AW, Oluka GK, Kato L, Ubaldo B, Kibengo F, Katende JS, Gombe B, Baine C, Odoch G, Mugaba S, Sande OJ; COVID-19 Immunoprofiling Team; Kaleebu P, Serwanga J. Pre-pandemic SARS-CoV-2-specific IFN-γ and antibody responses were low in Ugandan samples and significantly reduced in HIV-positive specimens. Front Immunol. 2023 Apr 19;14:1148877.
26. Aissatou A, Fokam J, Semengue ENJ, Takou D, Ka'e AC, Ambe CC, Nka AD, Djupsa SC, Beloumou G, Ciaffi L, Tchouaket MCT, Nayang ARM, Pabo WLT, Essomba RG, Halle EGE, Okomo MC, Bissek AZ, Leke R, Boum Y 2nd, Mballa GAE, Montesano C, Perno CF, Colizzi V, Ndjolo A. Pre-existing immunity to SARS-CoV-2 before the COVID-19 pandemic era in Cameroon: A comparative analysis according to HIV-status. Front Immunol. 2023 Mar 8;14:1155855.

**Supplementary Table 1: Joanna Briggs risk of bias assessment**

|  | CRITERION | Scoring of studies |
| --- | --- | --- |
| 1 | Was the sample representative of the target population? | Not applicable (no datasets had been collected with prior anticipation to be used for this pandemic-related question) |
| 2 | Were study participants recruited in an appropriate way? | Unclear in all studies |
| 3 | Was the sample size adequate? | Yes for Woodford, Baker, Souris (except for Senegal), Gelanew, Naidoo (N antibodies), Li (Nigeria, Uganda, Kenya) and No for the other studies or subsets (setting a threshold of having at least N=306 samples for 4% precision at 95% CI with expected proportion of positivity being 15%) |
| 4 | Were the study subjects and the setting described in detail? | No for Pedersen, Gelanew, and Gdoura and Yes for the other 15 studies if lenient about required information; most studies however did not give in-depth details |
| 5 | Was the data analysis conducted with sufficient coverage of the identified sample? | Yes for all studies, given that samples could be measured for antibodies with no/few missing measurements (although one cannot be certain of missingness at earlier stages of sampling) |
| 6 | Were objective, standard criteria used for the measurement of the condition? | Yes for all studies (based on providing definitions for positivity that are standard or defendable) |
| 7 | Was the condition measured reliably? | Unclear in all studies (it cannot be verified that antibody assays were performed reliably) |
| 8 | Was there appropriate statistical analysis? | Yes for all studies (if one requires only provision of positive and tested, not all studies gave confidence intervals, but these can be calculated in number of positives and of tested are given)) |
| 9 | Are all important confounding factors/subgroups/differences identified and accounted for? | No for all studies, although several did explore some factors (as delineated in Table 2) |
| 10 | Were subpopulations identified using objective criteria? | Yes, for subpopulations listed in Table 2 |

**Supplementary Table 2. Data on positivity and covariates of interest extracted**

| Author | Country | Antigenic targets | Antigen | Antibody | HIV high positivity | Malaria high burden | Dengue high burden | Positive | Tested |
| --- | --- | --- | --- | --- | --- | --- | --- | --- | --- |
| Pedersen | Gabon | S | S | IgG | No<10% | Yes | Yes | 12 | 116 |
| Pedersen | Gabon | N | N | IgG | No<10% | Yes | Yes | 20 | 116 |
| Pedersen | Senegal | S | S | IgG | No<10% | Yes | Yes | 20 | 144 |
| Pedersen | Senegal | N | N | IgG | No<10% | Yes | Yes | 14 | 144 |
| Borrega | Sierra Leone | N | N | IgG | No<10% | Yes | Yes | 52 | 120 |
| Borrega | Sierra Leone | RBD | RBD | IgG | No<10% | Yes | Yes | 72 | 120 |
| Borrega | Sierra Leone | S2 | S2 | IgG | No<10% | Yes | Yes | 42 | 120 |
| Tso | Tanzania | N | N | IgG | No<10% | No | Yes | 18 | 105 |
| Tso | Tanzania | S | S | IgG | No<10% | No | Yes | 3 | 105 |
| Tso | Zambia | N | N | IgG | Yes>10% | Yes | No | 13 | 99 |
| Tso | Zambia | S | S | IgG | Yes>10% | Yes | No | 4 | 99 |
| Emmerich | Magadascar | S1 Euroimmun | S1 | IgG | No<10% | No | Yes | 3 | 167 (0 b) |
| Emmerich | Magadascar | Nmod Euroimmun | N | IgG | No<10% | No | Yes | 2 | 167 (0 b) |
| Emmerich | Magadascar | N EDI | N | IgG | No<10% | No | Yes | 11 | 167 (10 b) |
| Emmerich | Magadascar | N Mikrogen | N | IgG | No<10% | No | Yes | 1 | 167 (3 b) |
| Emmerich | Ghana 1 | S1 Euroimmun | S1 | IgG | No<10% | Yes | Yes | 12 | 150 (1 b) |
| Emmerich | Ghana 1 | Nmod Euroimmun | N | IgG | No<10% | Yes | Yes | 14 | 150 (11 b) |
| Emmerich | Ghana 1 | N EDI | N | IgG | No<10% | Yes | Yes | 24 | 150 (17 b) |
| Emmerich | Ghana 1 | N Mikrogen | N | IgG | No<10% | Yes | Yes | 7 | 150 (7 b) |
| Emmerich | Ghana 2 | S1 Euroimmun | S1 | IgG | No<10% | Yes | Yes | 4 | 133 (3 b) |
| Emmerich | Ghana 2 | Nmod Euroimmun | N | IgG | No<10% | Yes | Yes | 34 | 133 (21 b) |
| Emmerich | Ghana 2 | N EDI | N | IgG | No<10% | Yes | Yes | 39 | 133 (15 b) |
| Emmerich | Ghana 2 | N Mikrogen | N | IgG | No<10% | Yes | Yes | 22 | 133 (10 b) |
| Emmerich | Nigeria | S1 Euroimmun | S1 | IgG | No<10% | Yes | Yes | 14 | 150 (7 b) |
| Emmerich | Nigeria | Nmod Euroimmun | N | IgG | No<10% | Yes | Yes | 42 | 150 (18 b) |
| Emmerich | Nigeria | N EDI | N | IgG | No<10% | Yes | Yes | 91 | 150 (19 b) |
| Emmerich | Nigeria | N Mikrogen | N | IgG | No<10% | Yes | Yes | 26 | 150 (6 b) |
| Yadouleton | Benin | S1 (Euroimmun) | S1 | IgG | No<10% | Yes | No | 8 | 60 (1b) |
| Yadouleton | Benin | S1 IgA (Euroimmun) | S1 | IgA | No<10% | Yes | No | 7 | 60 |
| Yadouleton | Benin | S1 (InBios) | S1 | IgG | No<10% | Yes | No | 1 | 60 |
| Yadouleton | Benin | N (Euroimmun) | N | IgG | No<10% | Yes | No | 8 | 60 (1b) |
| Nzoghe | Gabon | N all subclasses | N | All Ig | No<10% | Yes | Yes | 32 | 135 |
| Woodford | Mali | S | S | IgG | No<10% | Yes | Yes | 21 | 312 |
| Woodford | Mali | N | N | IgG | No<10% | Yes | Yes | 116 | 233 |
| Woodford | Mali | RBD | RBD | IgG | No<10% | Yes | Yes | 73 | 312 |
| Baker | Uganda | Spike RBD IgM | RBD | IgM | No<10% | Yes | No | 31 | 1077 |
| Baker | Uganda | Spike RBD IgG | RBD | IgG | No<10% | Yes | No | 7 | 1077 |
| Souris | DR Congo 1+2 | S1 | S1 | IgG | No<10% | Yes | Yes | 110 | 574 |
| Souris | DR Congo 1+2 | S2 | S2 | IgG | No<10% | Yes | Yes | 45 | 574 |
| Souris | DR Congo 1+2 | S1-RBD | RBD | IgG | No<10% | Yes | Yes | 33 | 574 |
| Souris | DR Congo 1+2 | S1-NTD | NTD | IgG | No<10% | Yes | Yes | 32 | 574 |
| Souris | DR Congo 1+2 | N | N | IgG | No<10% | Yes | Yes | 42 | 574 |
| Souris | Cameroon | S1 | S1 | IgG | Yes>10% | Yes | Yes | 124 | 383 |
| Souris | Cameroon | S2 | S2 | IgG | Yes>10% | Yes | Yes | 69 | 383 |
| Souris | Cameroon | S1-RBD | RBD | IgG | Yes>10% | Yes | Yes | 85 | 383 |
| Souris | Cameroon | S1-NTD | NTD | IgG | Yes>10% | Yes | Yes | 23 | 383 |
| Souris | Cameroon | N | N | IgG | Yes>10% | Yes | Yes | 22 | 383 |
| Souris | R Congo | S1 | S1 | IgG | No<10% | Yes | Yes | 73 | 536 |
| Souris | R Congo | S2 | S2 | IgG | No<10% | Yes | Yes | 58 | 536 |
| Souris | R Congo | S1-RBD | RBD | IgG | No<10% | Yes | Yes | 47 | 536 |
| Souris | R Congo | S1-NTD | NTD | IgG | No<10% | Yes | Yes | 28 | 536 |
| Souris | R Congo | N | N | IgG | No<10% | Yes | Yes | 41 | 536 |
| Souris | Senegal | S1 | S1 | IgG | No<10% | Yes | Yes | 27 | 162 |
| Souris | Senegal | S2 | S2 | IgG | No<10% | Yes | Yes | 6 | 162 |
| Souris | Senegal | S1-RBD | RBD | IgG | No<10% | Yes | Yes | 18 | 162 |
| Souris | Senegal | S1-NTD | NTD | IgG | No<10% | Yes | Yes | 8 | 162 |
| Souris | Senegal | N | N | IgG | No<10% | Yes | Yes | 7 | 162 |
| Ige | Nigeria | S1 Abbott | S1 | IgG | Yes>10% | Yes | Yes | 0 | 100 |
| Ige | Nigeria | N Eurommun | N | IgG | Yes>10% | Yes | Yes | 3 | 100 |
| Ige | Nigeria | S Eurommun | S | IgG | Yes>10% | Yes | Yes | 0 | 100 |
| Ige | Nigeria | N or S2 Mologic | N+S2 | IgG | Yes>10% | Yes | Yes | 16 | 100 |
| Traore | Mali | S | S | IgG | No<10% | Yes | Yes | 62 | 283 |
| Traore | Mali | RBD | RBD | IgG | No<10% | Yes | Yes | 19 | 283 |
| Traore | Mali | RBM | RBM | IgG | No<10% | Yes | Yes | 25 | 283 |
| Ingoba | R Congo | Virotech | N | IgG | No<10% | Yes | Yes | 3 | 82 |
| Diagne | Senegal | N or S2 | N+S2 | IgG | No<10% | Yes | Yes | 88 | 272 |
| Diagne | Senegal | N | N | IgG | No<10% | Yes | Yes | 32 | 152 |
| Yansouni | Senegal | N (Abbott Architect) | N | IgG | No<10% | Yes | Yes | 8 | 90 |
| Yansouni | Senegal | N IgM (Standard Q-SD) | N | IgM | No<10% | Yes | Yes | 9 | 100 |
| Yansouni | Senegal | N IgG (Standard Q-SD) | N | IgG | No<10% | Yes | Yes | 1 | 100 |
| Iriemenam | Nigeria | Tetracore RBD | RBD | IgG | Yes>10% | Yes | Yes | 2 | 99 |
| Iriemenam | Nigeria | Tetracore N | N | IgG | Yes>10% | Yes | Yes | 8 | 99 |
| Iriemenam | Nigeria | Tetracore RBD-N | RBD+N | IgG | Yes>10% | Yes | Yes | 1 | 99 |
| Iriemenam | Nigeria | SARS2MBA S | S | IgG | Yes>10% | Yes | Yes | 1 | 98 |
| Iriemenam | Nigeria | SARS2MBA RBD541 | RBD | IgG | Yes>10% | Yes | Yes | 3 | 98 |
| Iriemenam | Nigeria | SARS2MBA RBD591 | RBD | IgG | Yes>10% | Yes | Yes | 3 | 98 |
| Iriemenam | Nigeria | SARS2MBA N | N | IgG | Yes>10% | Yes | Yes | 14 | 98 |
| Iriemenam | Nigeria | xMAP N | N | IgG | Yes>10% | Yes | Yes | 7 | 100 |
| Iriemenam | Nigeria | xMAP RBD | RBD | IgG | Yes>10% | Yes | Yes | 2 | 100 |
| Iriemenam | Nigeria | xMAP S1 | S1 | IgG | Yes>10% | Yes | Yes | 0 | 100 |
| Iriemenam | Nigeria | RightSign S RBD | S | IgG/IgM | Yes>10% | Yes | Yes | 0 | 100 |
| Iriemenam | Nigeria | Tetracore RBD | RBD | IgG | Yes>10% | Yes | Yes | 11 | 200 |
| Iriemenam | Nigeria | Tetracore N | N | IgG | Yes>10% | Yes | Yes | 44 | 200 |
| Iriemenam | Nigeria | Tetracore RBD-N | RBD+N | IgG | Yes>10% | Yes | Yes | 12 | 200 |
| Iriemenam | Nigeria | SARS2MBA S | S | IgG | Yes>10% | Yes | Yes | 1 | 198 |
| Iriemenam | Nigeria | SARS2MBA RBD541 | RBD | IgG | Yes>10% | Yes | Yes | 11 | 198 |
| Iriemenam | Nigeria | SARS2MBA RBD591 | RBD | IgG | Yes>10% | Yes | Yes | 8 | 198 |
| Iriemenam | Nigeria | SARS2MBA N | N | IgG | Yes>10% | Yes | Yes | 8 | 198 |
| Iriemenam | Nigeria | xMAP N | N | IgG | Yes>10% | Yes | Yes | 30 | 200 |
| Iriemenam | Nigeria | xMAP RBD | RBD | IgG | Yes>10% | Yes | Yes | 8 | 200 |
| Iriemenam | Nigeria | xMAP S1 | S1 | IgG | Yes>10% | Yes | Yes | 0 | 200 |
| Iriemenam | Nigeria | RightSign S | S | IgG/IgM | Yes>10% | Yes | Yes | 0 | 200 |
| Steinhardt | Nigeria | N (Euroimmun) | N | IgG | Yes>10% | Yes | Yes | 38 | 213 (7b) |
| Steinhardt | Nigeria | N (Abbott) | N | IgG | Yes>10% | Yes | Yes | 13 | 212 |
| Gelanew | Ethiopia | RBD | RBD | IgG | No<10% | No | Yes | 30 | 365 |
| Gebrecherkos | Ethiopia | LFIA-Canea, (N and S) | N+S | IgM | No<10% | No | Yes | 0 | 50 |
| Gebrecherkos | Ethiopia | LFIA-Canea, (N and S) | N+S | IgG | No<10% | No | Yes | 0 | 50 |
| Gebrecherkos | Ethiopia | LFIA-Canea, (N and S) | N+S | IgG/IgM | No<10% | No | Yes | 0 | 50 |
| Gebrecherkos | Ethiopia | LFIA Cellex (N and S) | N+S | IgM | No<10% | No | Yes | 0 | 50 |
| Gebrecherkos | Ethiopia | LFIA Cellex (N and S) | N+S | IgG | No<10% | No | Yes | 0 | 50 |
| Gebrecherkos | Ethiopia | LFIA Cellex (N and S) (N and S) | N+S | IgG/IgM | No<10% | No | Yes | 0 | 50 |
| Gebrecherkos | Ethiopia | LFIA-VivaCheck (N and S) | N+S | IgM | No<10% | No | Yes | 0 | 50 |
| Gebrecherkos | Ethiopia | LFIA-VivaCheck (N and S) | N+S | IgG | No<10% | No | Yes | 0 | 50 |
| Gebrecherkos | Ethiopia | LFIA-VivaCheck (N and S) | N+S | IgG/IgM | No<10% | No | Yes | 0 | 50 |
| Gebrecherkos | Ethiopia | LFIA-Innovita (N and S) | N+S | IgM | No<10% | No | Yes | 2 | 50 |
| Gebrecherkos | Ethiopia | LFIA-Innovita (N and S) | N+S | IgG | No<10% | No | Yes | 0 | 50 |
| Gebrecherkos | Ethiopia | LFIA-Innovita (N and S) | N+S | IgG/IgM | No<10% | No | Yes | 2 | 50 |
| Gebrecherkos | Ethiopia | ECLIA Roche (N) | N | All Ig | No<10% | No | Yes | 0 | 50 |
| Mboumba Bouassa | Central Africa Republic | BIOSYNEX IgG | RBD | IgG | Yes>10% | Yes | No | 0 | 100 |
| Mboumba Bouassa | Central Africa Republic | BIOSYNEX IgM | RBD | IgM | Yes>10% | Yes | No | 3 | 100 |
| Mboumba Bouassa | Central Africa Republic | SIENNA IgG | RBD | IgG | Yes>10% | Yes | No | 0 | 100 |
| Mboumba Bouassa | Central Africa Republic | SIENNA IgM | RBD | IgM | Yes>10% | Yes | No | 8 | 100 |
| Mboumba Bouassa | Central Africa Republic | NG-test IgG | Unknown | IgG | Yes>10% | Yes | No | 1 | 95 |
| Mboumba Bouassa | Central Africa Republic | NG-test IgM | Unknown | IgM | Yes>10% | Yes | No | 9 | 95 |
| Vanroye | Diverse countries | Toda IgG/IgM (N) | N | IgG/IgM | Unknown | Yes | No | 6 | 195 |
| Vanroye | Diverse countries | Cellex IgG/IgM (N+S) | N+S | IgG/IgM | Unknown | Yes | No | 7 | 195 |
| Vanroye | Diverse countries | Mutli-G IgG/IgM (N+RBD+S) | N+RBD+S | IgG/IgM | Unknown | Yes | No | 13 | 195 |
| Vanroye | Diverse countries | Sure Screen IgG/IgM (N+RBD) | N+RBD | IgG/IgM | Unknown | Yes | No | 14 | 195 |
| Vanroye | Diverse countries | Strong Strep IgG/IgM (N+S) | N+S | IgG/IgM | Unknown | Yes | No | 17 | 195 |
| Vanroye | Diverse countries | QuickZen IgG/IgM (RBD) | RBD | IgG/IgM | Unknown | Yes | No | 24 | 195 |
| Vanroye | Diverse countries | Biohit IgG/IgM (N) | N | IgG/IgM | Unknown | Yes | No | 31 | 195 |
| Vanroye | Diverse countries | Singuway IgG/IgM (N+RBD) | N+RBD | IgG/IgM | Unknown | Yes | No | 41 | 195 |
| Vanroye | Diverse countries | Panbio IgG/IgM (N) | N | IgG/IgM | Unknown | Yes | No | 44 | 195 |
| Vanroye | Diverse countries | Dynamiker IgG/IgM (N+S) | N+S | IgG/IgM | Unknown | Yes | No | 57 | 195 |
| Vanroye | Diverse countries | Healgen IgG/IgM (S) | S | IgG/IgM | Unknown | Yes | No | 96 | 195 |
| Vanroye | Diverse countries | Wanti (ND) (S) | S | Unclear | Unknown | Yes | No | 78 | 195 |
| Vanroye | Diverse countries | Boson IgG/IgM (N) | N | IgG/IgM | Unknown | Yes | No | 100 | 195 |
| Lapidus | Cameroon | S1 | S1 | IgG | No<10% | Yes | Yes | 4 | 19 |
| Lapidus | Cameroon | S1 | S1 | IgM | No<10% | Yes | Yes | 2 | 19 |
| Lapidus | Senegal1 | S1 | S1 | IgG | No<10% | Yes | Yes | 53 | 120 |
| Lapidus | Senegal1 | S1 | S1 | IgM | No<10% | Yes | Yes | 40 | 120 |
| Lapidus | Senegal2 | S1 | S1 | IgG | No<10% | Yes | Yes | 45 | 67 |
| Lapidus | Senegal2 | S1 | S1 | IgM | No<10% | Yes | Yes | 56 | 67 |
| Lapidus | BurkinaFasso1 | S1 | S1 | IgG | No<10% | Yes | Yes | 21 | 88 |
| Lapidus | BurkinaFasso1 | S1 | S1 | IgM | No<10% | Yes | Yes | 10 | 88 |
| Lapidus | BurkinaFasso2 | S1 | S1 | IgG | No<10% | Yes | Yes | 10 | 25 |
| Lapidus | BurkinaFasso2 | S1 | S1 | IgM | No<10% | Yes | Yes | 12 | 25 |
| Lapidus | Ghana | S1 | S1 | IgG | No<10% | Yes | Yes | 37 | 45 |
| Lapidus | Ghana | S1 | S1 | IgM | No<10% | Yes | Yes | 21 | 45 |
| Gdoura | Tunisia | Vidas S1 RBD | RBD | IgM | No<10% | No | No | 7 | 119 |
| Gdoura | Tunisia | Vidas S1 RBD | RBD | IgG | No<10% | No | No | 0 | 119 |
| Gdoura | Tunisia | Mindray S+N | S+N | IgM | No<10% | No | No | 3 | 119 |
| Gdoura | Tunisia | Mindray S+N | S+N | IgG | No<10% | No | No | 5 | 119 |
| Gdoura | Tunisia | Cobas N | N | All Ig | No<10% | No | No | 0 | 119 |
| Gdoura | Tunisia | Access S1 RBD | RBD | IgG | No<10% | No | No | 0 | 119 |
| Naidoo | South Africa | Elecsys N | N | All Ig | Yes>10% | No | No | 1 | 316 |
| Naidoo | South Africa | Elecsyn S | S | All Ig | Yes>10% | No | No | 0 | 194 |
| Li | Nigeria | S2 | S2 | IgG | Yes>10% | Yes | Yes | 9 | 361 |
| Li | Uganda | S2 | S2 | IgG | Yes>10% | Yes | No | 49 | 399 |
| Li | Tanzania | S2 | S2 | IgG | Yes>10% | No | Yes | 13 | 234 |
| Li | Kenya | S2 | S2 | IgG | Yes>10% | No | Yes | 46 | 653 |
| Nantambi | Uganda | N | N | IgG | Yes>10% | Yes | No | 17 | 110 |
| Nantambi | Uganda | S | S | IgG | Yes>10% | Yes | No | 3 | 110 |
| Aissatou | Cameroon | N | N | IgG | Yes>10% | Yes | Yes | 21 | 288 |
| Aissatou | Cameroon | N | N | IgM | Yes>10% | Yes | Yes | 21 | 288 |
| Aissatou | Cameroon | N | N | IgG/IgM | Yes>10% | Yes | Yes | 39 | 288 |

**Supplementary Table 3. Data on paired IgM and IgG measurements on the same samples with the same assay**

| AUTHOR | COUNTRY | IgM only | IgG only | Both | None |
| --- | --- | --- | --- | --- | --- |
| Baker | Uganda | 31 | 5 | 2 | 1039 |
| Gebrecherkos | Ethiopia 1 | 0 | 0 | 0 | 50 |
| Gebrecherkos | Ethiopia 2 | 0 | 0 | 0 | 50 |
| Gebrecherkos | Ethiopia 3 | 0 | 0 | 0 | 50 |
| Gebrecherkos | Ethiopia 4 | 2 | 0 | 0 | 48 |
| Mboumpa Bouassa | Central Afr Rep 1 | 3 | 0 | 0 | 97 |
| Mboumpa Bouassa | Central Afr Rep 2 | 8 | 0 | 0 | 92 |
| Lapidus | Cameroon | 0 | 2 | 2 | 15 |
| Lapidus | Senegal 1 | 7 | 20 | 33 | 60 |
| Lapidus | Senegal 2 | 17 | 6 | 39 | 5 |
| Lapidus | Burkina Faso 1 | 3 | 14 | 7 | 64 |
| Lapidus | Burkina Faso 2 | 5 | 3 | 7 | 10 |
| Lapidus | Ghana | 3 | 19 | 18 | 5 |
| Gdoura | Tunisia (Vidas) | 7 | 0 | 0 | 112 |
| Gdoura | Tunisia (Mindray) | 2 | 4 | 1 | 112 |
| Aissatou | Cameroon | 18 | 18 | 3 | 249 |

**Supplementary Table 4. Data on *Plasmodium* parasitemia (Par) and anti-SARS-CoV-2 antibodies (Ab)**

| AUTHOR | Ab+Par+ | Ab-Par+ | Ab+/Par- | Ab-Par- |
| --- | --- | --- | --- | --- |
| Emmerich 1 | 9 | 46 | 5 | 90 |
| Emmerich 2 | 6 | 49 | 6 | 89 |
| Emmerich 3 | 12 | 43 | 12 | 83 |
| Emmerich 4 | 1 | 54 | 6 | 89 |

The respective p-values are 0.032, 0.32, 0.14, and 0.24 with odds ratios >1 in the first 3 and odds ratio <1 in the fourth.

Besides these data, information on Plasmodium parasitemia and anti-SARS-C0V-2 antibodies was provided by Traore et al.; the correlation coefficients were 0.10 (p=0.09), 0.06 (p=0.35), and -0.07 (p=0.27) against the readings of optical density for anti-S, anti-RBD, and anti-RBM antibodies, respectively. Also Steinhardt et al. assessed 4 Plasmodium antigens and their levels were not significantly different in patients with and without SARS-CoV-2 cross-reactive Euroimmun or Abbott antibodies; nevertheless, median values for all 4 antigens (HRP2, PvLDH, pAldolase, pLDH) were always higher in patients with versus those without SARS-CoV-2 cross-reactivity (p=0.17, p=0.97, p=0.21, and p=0.16 for Euroimmun and p=0.34, p=0.39, p=0.24, and p=0.12 for Abbott).

Therefore, of the 15 assessments, 13 are in the direction of higher parasitemia associated with SARS-CoV-2 cross-reactivity and 2 are in the opposite direction (inverse association). A meta-analysis of the 15 p-values for these data (converted to one-tailed, which accounts that 2 of the 15 have inverse associations) using the Fisher method yields summary p=0.0009. Alternatively, meta-analysis using the additive Edgington method yields summary p=0.00012 and meta-analysis using the normal Edgington method yields summary p=0.00018.

**Supplementary Table 5. Data on HIV status and anti-SARS-CoV-2 antibodies (Ab)**

| AUTHOR | Ab+HIV+ | Ab-HIV+ | Ab+HIV- | Ab-HIV- |
| --- | --- | --- | --- | --- |
| Tso Tanzania | 0 | 7 | 18 | 80 |
| Tso Zambia | 4 | 39 | 9 | 47 |
| Baker Uganda | 17 | 442 | 21 | 597 |
| Nantambi N | 3 | 33 | 14 | 54 |
| Nantambi S | 2 | 35 | 1 | 67 |
| Aissatou IgG | 10 | 153 | 11 | 114 |
| Aissatou IgM | 9 | 154 | 12 | 113 |
| Aissatou IgG/IgM | 18 | 145 | 21 | 104 |

**Supplementary Excluded References – Excluded studies [with reason for exclusion]**

1. Lutalo T, Nalumansi A, Olara D, Kayiwa J, Ogwang B, Odwilo E, Watera C, Balinandi S, Kiconco J, Nakaseegu J, Serwanga J, Kikaire B, Ssemwanga D, Abiko B, Nsereko C, Cotten M, Buule J, Lutwama J, Downing R, Kaleebu P. Evaluation of the performance of 25 SARS-CoV-2 serological rapid diagnostic tests using a reference panel of plasma specimens at the Uganda Virus Research Institute. Int J Infect Dis. 2021 Nov;112:281-287. [Excluded pre-pandemic samples positive by other assays]
2. Matefo L, Cloete VV, Armand BP, Dominique G, Samantha P, John F, Craig T, Daniel W, Theresa L, Sunetra G, Maréza B, Danelle VJ, Jane BF. Validation of laboratory developed serology assays for detection of IgG antibody to severe acute respiratory syndrome coronavirus 2 in the South African population. J Virol Methods. 2022 Sep;307:114571. [Calibration]
3. Nyagwange J, Kutima B, Mwai K, Karanja HK, Gitonga JN, Mugo D, Uyoga S, Tuju J, Ochola-Oyier LI, Ndungu F, Bejon P, Agweyu A, Adetifa IMO, Scott JAG, Warimwe GM. Comparative performance of WANTAI ELISA for total immunoglobulin to receptor binding protein and an ELISA for IgG to spike protein in detecting SARS-CoV-2 antibodies in Kenyan populations. J Clin Virol. 2022 Jan;146:105061. [Assays previously developed with calibration on African samples]
4. Wiens KE, Mawien PN, Rumunu J, et al. Seroprevalence of Severe Acute Respiratory Syndrome Coronavirus 2 IgG in Juba, South Sudan, 2020(1). Emerg Infect Dis 2021; 27(6): 1598-606. [Calibration]
5. Sisay A, Tesfaye A, Desale A, Ataro I, Woldesenbet Z, Nigusse B, Tayachew A, Kebede A, Desta AF. Diagnostic Performance of SARS-CoV-2 IgM/IgG Rapid Test Kits for the Detection of the Novel Coronavirus in Ethiopia. J Multidiscip Healthc. 2021 Jan 27;14:171-180. [No pre-pandemic samples]
6. Stoddard CI, Sung K, Ojee E, Adhiambo J, Begnel ER, Slyker J, Gantt S, Matsen FA 4th, Kinuthia J, Wamalwa D, Overbaugh J, Lehman DA. Distinct Antibody Responses to Endemic Coronaviruses Pre- and Post-SARS-CoV-2 Infection in Kenyan Infants and Mothers. Viruses. 2022 Jul 12;14(7):1517. [Pre-pandemic samples cannot be separated]
7. Péré H, Mboumba Bouassa RS, Tonen-Wolyec S, Podglajen I, Veyer D, Bélec L. Analytical performances of five SARS-CoV-2 whole-blood finger-stick IgG-IgM combined antibody rapid tests. J Virol Methods. 2021 Apr;290:114067. [No African samples]
8. Tazi S, Kabbaj H, Zirar J, Zouaki A, El Amin G, El Himeur O, Seffar M. Comparative Performance Evaluation of FilmArray BioFire RP2.1 and MAScIR 2.0 Assays for SARS-CoV-2 Detection. Adv Virol. 2022 Jun 1;2022:4510900. [No pre-pandemic samples, nucleic acid assays evaluated]
9. Arinola OG, Edem VF, Rahamon SK, Yaqub SA, Fashina AO, Alonge TO. Sars-Cov-2 Infection Screening Using Two Serological Testing Methods. Niger J Physiol Sci. 2020 Dec 31;35(2):117-121. [No pre-pandemic samples]
10. Gededzha MP, Mampeule N, Jugwanth S, Zwane N, David A, Burgers WA, Blackburn JM, Grove JS, George JA, Sanne I, Scott L, Stevens W, Mayne ES. Performance of the EUROIMMUN Anti-SARS-CoV-2 ELISA Assay for detection of IgA and IgG antibodies in South Africa. PLoS One. 2021 Jun 23;16(6):e0252317. [Pre-pandemic samples cannot be separated]
11. Hussein NA, Ali EAA, El-Hakim AE, Tabll AA, El-Shershaby A, Salamony A, Shaheen MNF, Ali I, Elshall M, Shahein YE. Assessment of specific human antibodies against SARS-CoV-2 receptor binding domain by rapid in-house ELISA. Hum Antibodies. 2022;30(2):105-115. [No pre-pandemic samples]
12. Deschermeier C, Ehmen C, von Possel R, Murawski C, Rushton B, Amuasi J, Sarpong N, Maiga-Ascofaré O, Rakotozandrindrainy R, Asogun D, Ighodalo Y, Oestereich L, Duraffour S, Pahlmann M, Emmerich P. Fcγ-Receptor-Based Enzyme-Linked Immunosorbent Assays for Sensitive, Specific, and Persistent Detection of Anti-SARS-CoV-2 Nucleocapsid Protein IgG Antibodies in Human Sera. J Clin Microbiol. 2022 Jun 15;60(6):e0007522. [Calibration of new ELISAs; also includes data with commercial Euroimmun that are eligible, but are also included in Emmerich et al.]
13. Uyoga S, Adetifa IMO, Karanja HK, Nyagwange J, Tuju J, Wanjiku P, Aman R, Mwangangi M, Amoth P, Kasera K, Ng'ang'a W, Rombo C, Yegon C, Kithi K, Odhiambo E, Rotich T, Orgut I, Kihara S, Otiende M, Bottomley C, Mupe ZN, Kagucia EW, Gallagher KE, Etyang A, Voller S, Gitonga JN, Mugo D, Agoti CN, Otieno E, Ndwiga L, Lambe T, Wright D, Barasa E, Tsofa B, Bejon P, Ochola-Oyier LI, Agweyu A, Scott JAG, Warimwe GM. Seroprevalence of anti-SARS-CoV-2 IgG antibodies in Kenyan blood donors. Science. 2021 Jan 1;371(6524):79-82. [Calibration]
14. Crowell TA, Daud II, Maswai J, Owuoth J, Sing'oei V, Imbach M, Dear N, Sawe F, Eller LA, Polyak CS, Ake JA; AFRICOS Study Group. Severe acute respiratory syndrome coronavirus-2 antibody prevalence in people with and without HIV in rural Western Kenya, January to March 2020. AIDS. 2021 Nov 15;35(14):2401-2404. [No pre-pandemic samples]
15. Meinus C, Singer R, Nandi B, Jagot O, Becker-Ziaja B, Karo B, Mvula B, Jansen A, Baumann J, Schultz A. SARS-CoV-2 prevalence and immunity: a hospital-based study from Malawi. Int J Infect Dis. 2022 Mar;116:157-165. [Calibration]
16. Bonguili NCB, Fritz M, Lenguiya LH, Mayengue PI, Koukouikila-Koussounda F, Dossou-Yovo LR, Badzi CN, Leroy EM, Niama FR. Early Circulation of SARS-CoV-2, Congo, 2020. Emerg Infect Dis. 2022 Apr;28(4):878-880. [Calibration]
17. Serwanga J, Ankunda V, Sembera J, Kato L, Oluka GK, Baine C, Odoch G, Kayiwa J, Auma BO, Jjuuko M, Nsereko C, Cotten M, Onyachi N, Muwanga M, Lutalo T, Fox J, Musenero M, Kaleebu P; COVID-19 Immunoprofiling Team. Rapid, early, and potent Spike-directed IgG, IgM, and IgA distinguish asymptomatic from mildly symptomatic COVID-19 in Uganda, with IgG persisting for 28 months. Front Immunol. 2023 Mar 16;14:1152522. [Calibration]
18. Oluka GK, Namubiru P, Kato L, Ankunda V, Gombe B, Cotten M; COVID-19 Immunoprofiling Team; Musenero M, Kaleebu P, Fox J, Serwanga J. Optimisation and Validation of a conventional ELISA and cut-offs for detecting and quantifying anti-SARS-CoV-2 Spike, RBD, and Nucleoprotein IgG, IgM, and IgA antibodies in Uganda. Front Immunol. 2023 Mar 14;14:1113194. [Calibration]
19. Benabdessalem C, Hamouda WB, Marzouki S, Faye R, Mbow AA, Diouf B, Ndiaye O, Dia N, Faye O, Sall AA, Diagne CT, Amellal H, Ezzikouri S, Mioramalala DJN, Randrianarisaona F, Trabelsi K, Boumaiza M, Hamouda SB, Ouni R, Bchiri S, Chaaban A, Gdoura M, Gorgi Y, Sfar I, Yalaoui S, Khelil JB, Hamzaoui A, Abdallah M, Cherif Y, Petres S, Mok CKP, Escriou N, Quesney S, Dellagi K, Schoenhals M, Sarih M, Vigan-Womas I, Bettaieb J, Rourou S, Barbouche MR, Ahmed MB. Development and comparative evaluation of SARS-CoV-2 S-RBD and N based ELISA tests in various African endemic settings. Diagn Microbiol Infect Dis. 2023 Apr;105(4):115903. [Prepandemic samples cannot be separated]
20. Briggs J, Takahashi S, Nayebare P, Cuu G, Rek J, Zedi M, Kizza T, Arinaitwe E, Nankabirwa JI, Kamya M, Jagannathan P, Jacobson K, Rosenthal PJ, Dorsey G, Greenhouse B, Ssewanyana I, Rodríguez-Barraquer I. Seroprevalence of Antibodies to SARS-CoV-2 in Rural Households in Eastern Uganda, 2020-2022. JAMA Netw Open. 2023 Feb 1;6(2):e2255978. [Calibration]
21. Vigan-Womas I, Spadoni JL, Poiret T, Taïeb F, Randrianarisaona F, Faye R, Mbow AA, Gaye A, Dia N, Loucoubar C, Ny Mioramalala DJ, Ratovoson R, Randremanana RV, Sall AA, Seydi M, Noirel J, Moreau G, Simon A, Holenya P, Meyniel JP, Zagury JF, Schoenhals M. Linear epitope mapping of the humoral response against SARS-CoV-2 in two independent African cohorts. Sci Rep. 2023 Jan 16;13(1):782. [Pre-pandemic samples used as controls for peptide array analysis]
22. Nyagwange J, Kutima B, Mwai K, Karanja HK, Gitonga JN, Mugo D, Sein Y, Wright D, Omuoyo DO, Nyiro JU, Tuju J, Nokes DJ, Agweyu A, Bejon P, Ochola-Oyier LI, Scott JAG, Lambe T, Nduati E, Agoti C, Warimwe GM. Serum immunoglobulin G and mucosal immunoglobulin A antibodies from prepandemic samples collected in Kilifi, Kenya, neutralize SARS-CoV-2 in vitro. Int J Infect Dis. 2023 Feb;127:11-16. [Assay previously calibrated in African samples]
23. Gdoura M, Ghaloum FB, Hamida MB, Chamsa W, Triki H, Bahloul C. Development of an in-house quantitative ELISA for the evaluation of different Covid-19 vaccines in humans. Sci Rep. 2022 Jul 4;12(1):11298. doi: 10.1038/s41598-022-15378-1. [all relevant data already included in another article].
24. Iriemenam NC, Ige FA, Greby SM, et al. Validation of xMAP SARS-CoV-2 Multi-Antigen IgG assay in Nigeria. PLoS One. 2022 Apr 1;17(4):e0266184. [relevant data on 200/213 patients included in another article].

**Supplementary Figure: Meta-analysis of positivity rates for anti-SARS-COV-2 antibodies in pre-pandemic samples for studies from countries or settings with (A) high malaria burden and high dengue burden, (B) high malaria burden and low dengue burden, (C) low malaria burden and high dengue burden, and (D) low malaria burden and low dengue burden**

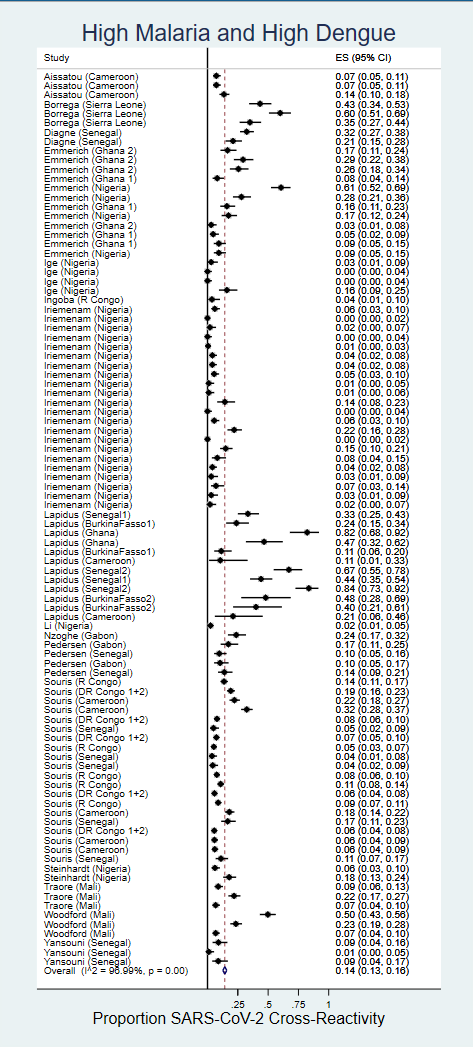

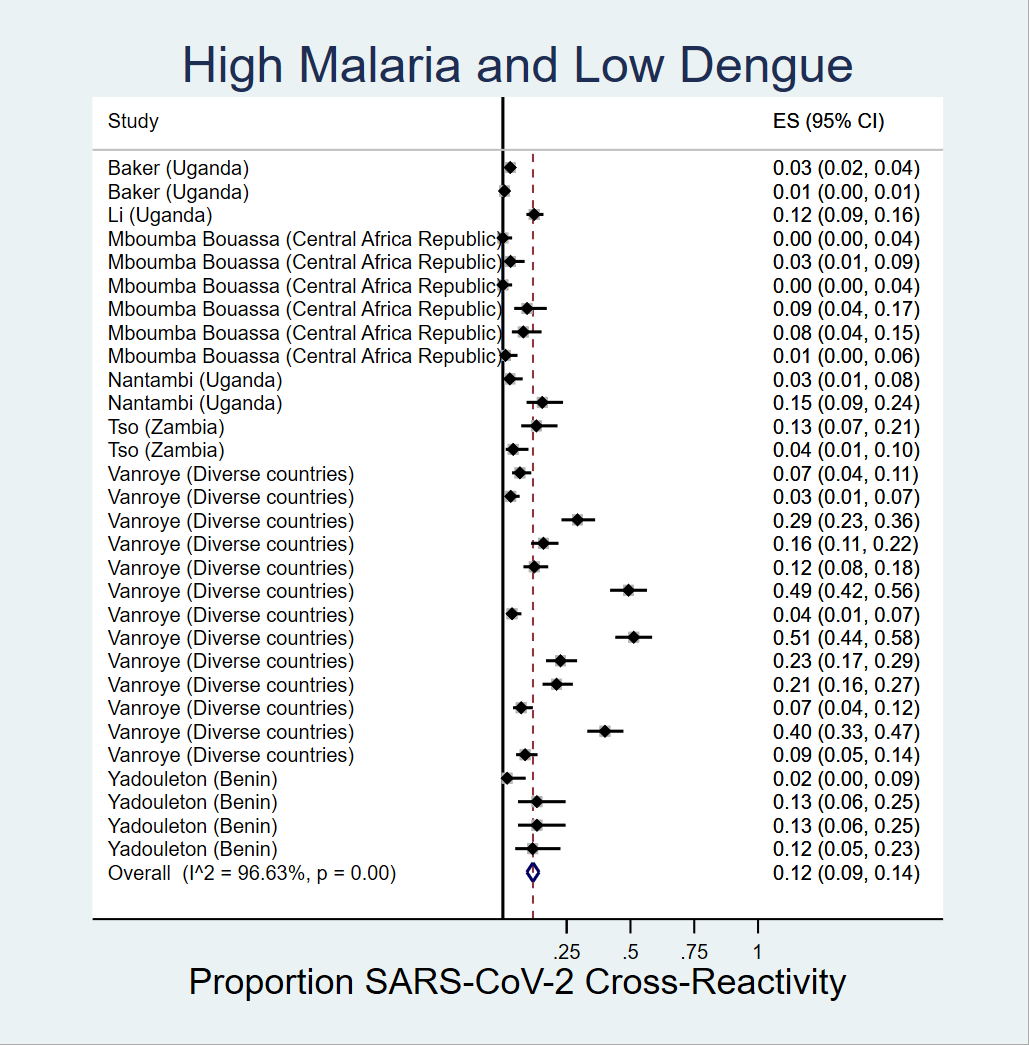

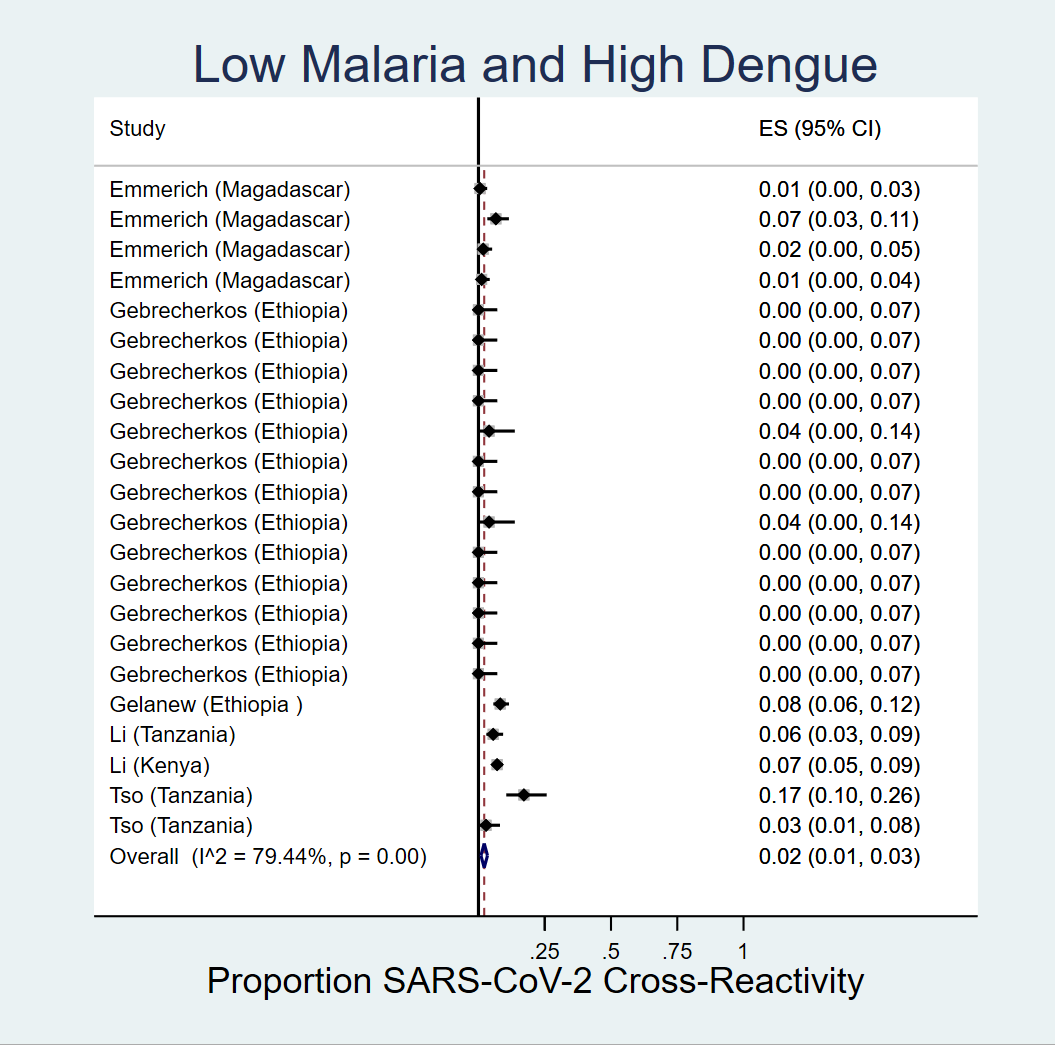

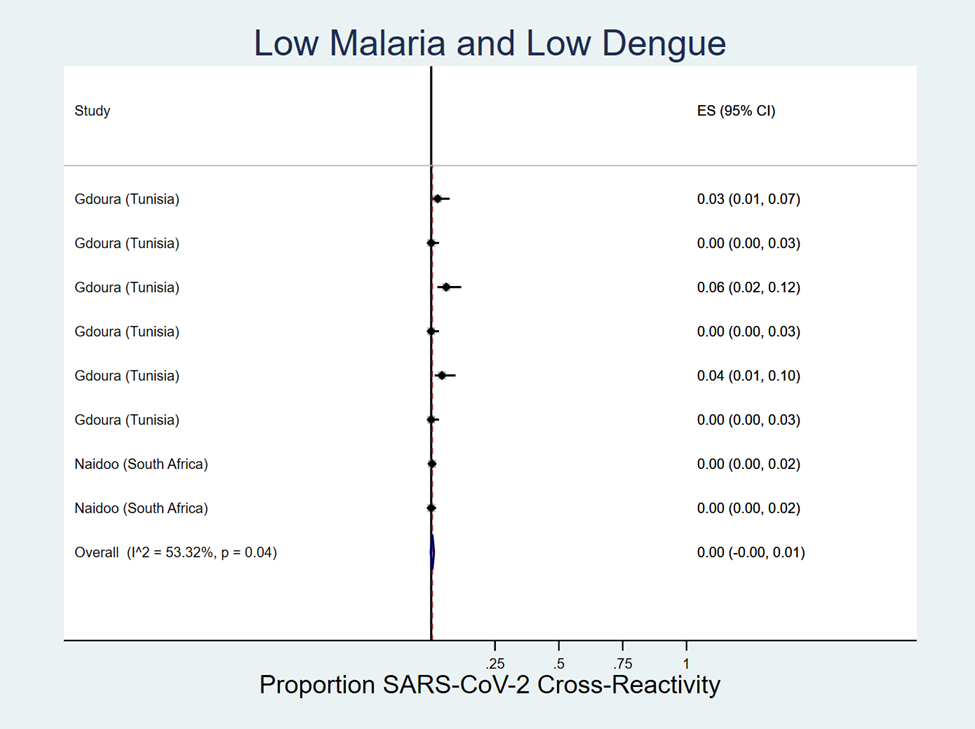
